# Transmission of SARS-CoV-2 from Children and Adolescents

**DOI:** 10.1101/2020.10.10.20210492

**Authors:** Victoria T. Chu, Anna R. Yousaf, Karen Chang, Noah G. Schwartz, Clinton J. McDaniel, Christine M. Szablewski, Marie Brown, Kathryn Winglee, Scott H. Lee, Zhaohui Cui, Adebola Adebayo, Tiffiany Aholou, Minal M. Amin, Peter Aryee, Cindy Castaneda, Trudy Chambers, Amy C. Fleshman, Christin Goodman, Tony Holmes, Asha Ivey-Stephenson, Emiko Kamitani, Susan Katz, Jennifer K. Knapp, Maureen Kolasa, Maranda Lumsden, Erin Mayweather, Asfia Mohammed, Anne Moorman, Alpa Patel-Larson, Lara Perinet, Mark Pilgard, Deirdre D. Pratt, Shanica Railey, Jaina Shah, Dawn Tuckey, Emilio Dirlikov, Dale A. Rose, Julie Villanueva, Alicia M. Fry, Aron J. Hall, Hannah L. Kirking, Jacqueline E. Tate, Cherie L. Drenzek, Tatiana M. Lanzieri, Rebekah J. Stewart

## Abstract

A better understanding of SARS-CoV-2 transmission from children and adolescents is crucial for informing public health mitigation strategies. We conducted a retrospective cohort study among household contacts of primary cases defined as children and adolescents aged 7⍰19 years with laboratory evidence of SARS-CoV-2 infection acquired during an overnight camp outbreak. Among household contacts, we defined secondary cases using the Council of State and Territorial Epidemiologists definition. Among 526 household contacts of 224 primary cases, 48 secondary cases were identified, corresponding to a secondary attack rate of 9% (95% confidence interval [CI], 7%–12%). Our findings show that children and adolescents can transmit SARS-CoV-2 to adult contacts and other children in a household setting.

## Introduction

Although children can experience severe illness from SARS-CoV-2 infection including multisystem inflammatory syndrome and death in rare cases,^1^ most experience mild or asymptomatic illness^2-4^ resulting in under-recognition of pediatric cases. Some studies suggest low secondary transmission from young children.^5-7^ However, closure of schools and other youth-centric settings early in the pandemic combined with mitigation measures and selective testing limit the reliability of these conclusions.^8,9^ Transmission to adults is of particular concern given the higher risk for severe illness from COVID-19 in older adults.^10,11^ Here we describe secondary attack rates (SAR) among household contacts of children and adolescents who acquired SARS-CoV-2 infection during an outbreak at an overnight camp in June 2020.^12^

## Methods

During July 17–August 24, 2020, the Centers for Disease Control and Prevention (CDC) collaborated with local and state health departments to contact all camp attendees and their parents or guardians for a phone interview. Using a structured questionnaire, we collected demographic and clinical characteristics of camp attendees, SARS-CoV-2 testing history, and a list of household contacts. We conducted a retrospective cohort study among household contacts exposed to camp attendees aged 7–19 years with self-reported evidence of SARS-CoV-2 infection by molecular or antigen testing. We interviewed each household contact to obtain dates of exposure to the camp attendee during their infectious period, as well as SARS-CoV-2 testing history, presence of COVID-19 symptoms, and potential community exposures.

COVID-19 cases among camp attendees were defined as those with self-reported laboratory evidence of SARS-CoV-2 infection by molecular or antigen testing. Among household contacts (i.e., persons who stayed ≥1 night in the household during the camp attendee case’s infectious period), COVID-19 cases and non-cases were categorized using the Council of State and Territorial Epidemiologists definitions approved on August 5, 2020.^13^ We did not distinguish between confirmed and probable cases among household contacts. Hereafter, we define a “primary case” as a camp attendee with the earliest onset date in the household and a “secondary case” as a household contact with confirmed or probable COVID-19. If multiple camp attendee cases resided in the same household, they were defined as coprimary cases.

We included households for which household contacts provided sufficient information for secondary case or non-case classification and excluded households in which the household contact case had a symptom onset date prior to or <2 days after the symptom onset date of the camp attendee. We described frequencies of categorical variables (i.e., demographic and clinical characteristics for primary and secondary cases, household interactions, and potential community exposures) and median, interquartile ranges (IQR), and ranges for quantitative variables (i.e., age). We used the Pearson’s chi-squared test and the Wilcoxon rank-sum test to compare the sex and age of household contacts who were interviewed with those who were not, respectively. We calculated the overall SAR by two approaches: 1) the percentage of secondary cases among all household contacts and 2) the percentage of secondary cases, excluding household contacts who were not tested for SARS-CoV-2. The 95% CI of these percentages was calculated using the Wilson score interval.

This activity was reviewed by CDC and the Georgia Department of Public Health and was conducted consistently with applicable federal laws and CDC policy as defined in 45 C.F.R. part 46, 21 C.F.R. part 56; 42 U.S.C. 241(d);⍰5 U.S.C. 552a; 44 U.S.C. 3501 et seq. For camp attendees aged <18 years, we obtained parental or guardian permission as well as verbal assent from the camp attendee.

## Results

We identified 224 primary cases in 194 households with 526 household contacts (Figure 1). In total, there were 163 households with one primary case and 456 household contacts, 30 households with two coprimary cases and 68 household contacts, and one household with three coprimary cases and two household contacts.

**Figure 1.**
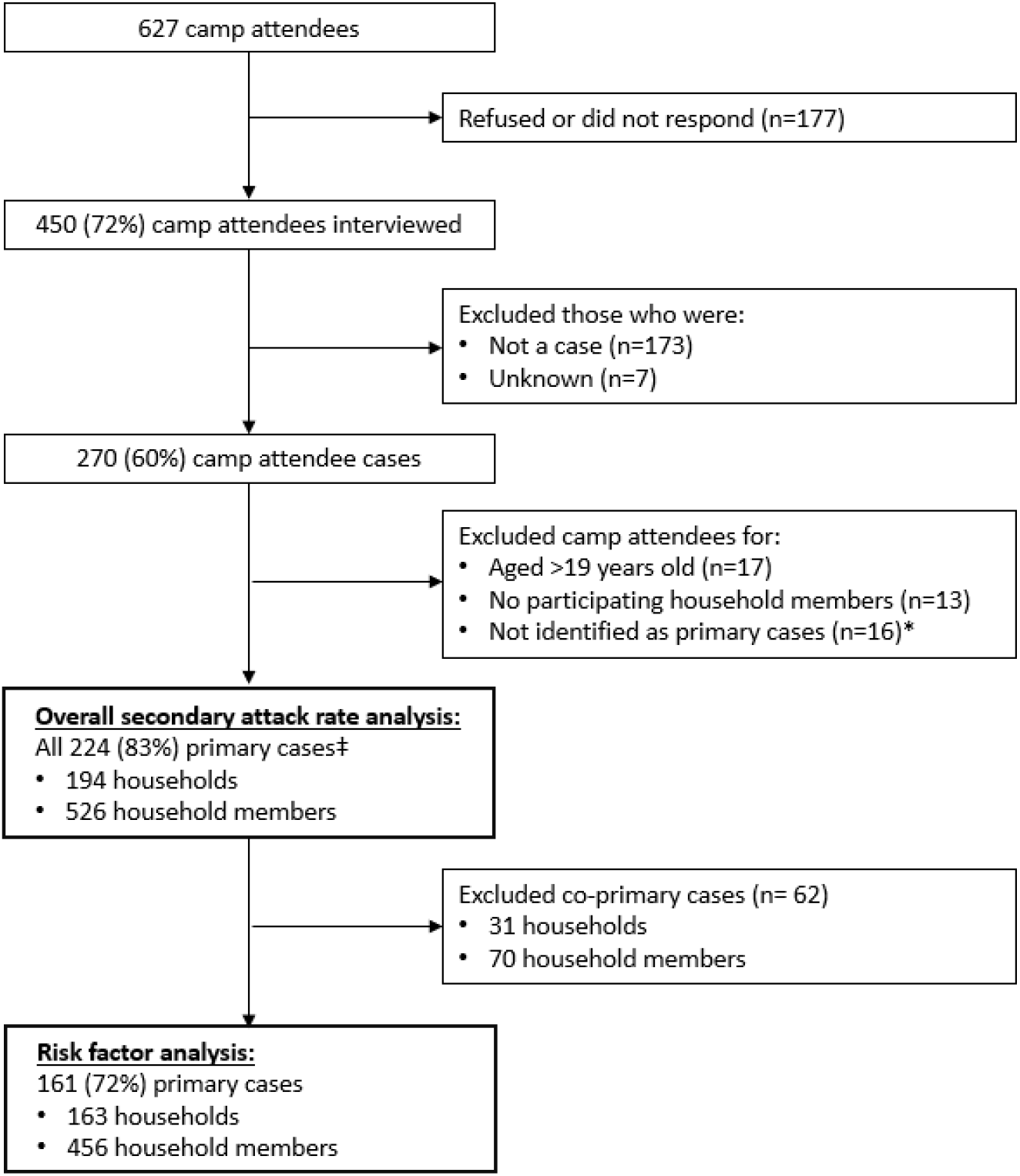
Flow diagram for enrollment of persons with COVID-19 identified as primary cases and their household contacts* *A primary case was defined as a camp attendee with self-reported evidence of SARS-CoV-2 infection by molecular or antigen testing and the earliest onset date in the household. A household contact was defined as a person who stayed ≥1 night in the household during the primary case’s infectious period (i.e., 2 days prior to the onset date until 10 days after the onset date). †Not identified as primary case includes 4 missing onset date, and 12 with household contacts with an onset date prior to or <2 days after the onset date of the camp attendee. ‡The primary case was defined as the case with the earliest onset date in the household.

The 224 primary cases had a median age of 14 years; 115 (51%) were female, and 198 (88%) were non-Hispanic White (Table 1). Of these, 184 (82%) were symptomatic, reporting constitutional symptoms (153; 68%), upper respiratory symptoms (110; 49%), new olfactory or taste disorders (64; 29%), gastrointestinal symptoms (49; 22%), or lower respiratory symptoms (34; 15%). None of the primary cases were hospitalized.

**Table 1.** Demographic and clinical characteristics of primary cases and their household contacts*

|  | **Primary Cases**<br>**n = 224** |  | **Household Contacts**<br>**n = 526<sup>†</sup>** |  |
| --- | --- | --- | --- | --- |
| **Characteristics** | **n** | **%** | **n** | **%** |
| Age (years): median (interquartile range) | 14 (7–19) |  | 46 (20–51) |  |
| Sex |  |  |  |  |
| Female | 115 | 51 | 262 | 50 |
| Male | 109 | 49 | 262 | 50 |
| Unknown | 0 | 0 | 2 | <1 |
| Race/ethnicity group |  |  |  |  |
| Non-Hispanic White | 198 | 88 | 400 | 76 |
| Non-Hispanic Black | 0 | 0 | 1 | <1 |
| Hispanic or Latino | 9 | 4 | 10 | 2 |
| Other | 6 | 3 | 10 | 2 |
| Unknown | 11 | 5 | 101 | 19 |
| ≥1 underlying medical condition<sup>‡</sup> | 14 | 6 | 74 | 14 |
| Chronic lung disease | 12 | 6 | 28 | 6 |
| Cardiovascular disease | 0 | 0 | 26 | 5 |
| Diabetes Mellitus | 0 | 0 | 10 | 2 |
| Immunocompromising condition or medication | 0 | 0 | 10 | 2 |
| Evidence of SARS-CoV-2 infection<sup>§</sup> |  |  |  |  |
| Positive | 224 | 100 | 46 | 9 |
| Negative | 0 | 0 | 331 | 63 |
| Never tested | 0 | 0 | 149 | 28 |

\*A primary case was defined as a camp attendee with self-reported evidence of SARS-CoV-2 infection by molecular or antigen testing and the earliest onset date in the household. A household contact was defined as a person who stayed ≥1 night in the household during the primary case's infectious period (i.e., 2 days prior to the onset date until 10 days after the onset date).
<sup>†</sup>Only age, sex, and SARS-CoV-2 testing status were known for the 80 household contacts who were not interviewed.
<sup>‡</sup>Denominator was 216 for primary cases and 441 for household contacts.
<sup>§</sup>A COVID-19 case was defined as laboratory evidence of SARS-CoV-2 infection included positive molecular or antigen testing and was self-reported.

The 526 household contacts had a median age of 46 years (range, 1–83); 262 (50%) were female (Table 1). Among the 526 household contacts, 351 (67%) were parents, 161 (31%) were siblings, 11 (2%) were extended family members, and 3 (1%) were non-familial contacts. Both the age and sex distribution of the household contacts who were interviewed (n = 446) and were not interviewed (n = 80) was similar (age: p-value = 0.68; sex: p-value = 0.90). Among 434 (84%) interviewed household contacts, 400 (90%) were non-Hispanic White, 74 (14%) had at least one underlying medical condition; among contacts aged ≥22 years, 92% (255/276) reported college education or higher.

We identified 48 household contacts as secondary cases (Table 2); 25 (52%) were male, and 44 (92%) were symptomatic (Table 2). None of the 7 secondary cases among contacts aged <18 years were hospitalized; 4 (10%) of 41 secondary cases among household contacts aged ≥18 years (aged 49–77 years) were hospitalized. Of these 4 hospitalized contacts, hospital length of stay varied from 5–11 days, and 2 (50%) had no underlying medical conditions.

**Table 2.** Demographic and clinical characteristics of household contacts with COVID-19 and other household contacts

|  | **Household Contacts with**<br>**COVID-19**<br>**n = 48** |  | **Other Household Contacts**<br>**n = 478\*** |  |
| --- | --- | --- | --- | --- |
| **Household contact**<br>**characteristics** | **n** | **%** | **N** | **%** |
| ≥1 underlying medical condition† | 12 | 26 | 63 | 13 |
| Chronic lung disease | 6 | 13 | 22 | 6 |
| Cardiovascular disease | 5 | 11 | 20 | 5 |
| Diabetes mellitus | 3 | 6 | 7 | 2 |
| Immunocompromising condition or medication | 4 | 9 | 6 | 2 |
| Evidence of SARS-CoV-2 infection‡ |  |  |  |  |
| Positive | 46 | 96 | 0 | 0 |
| Negative | 0 | 0 | 331 | 69 |
| Never tested | 2 | 4 | 147 | 31 |
| Frequency of SARS-CoV-2 testing§ |  |  |  |  |
| 0 | 3 | 6 | 162 | 34 |
| 1 | 34 | 71 | 227 | 47 |
| ≥2 | 11 | 23 | 89 | 19 |

\*Only SARS-CoV-2 testing status was known for the 80 household contacts who were not interviewed; none of whom were identified as household contacts with COVID-19.
†Denominator was 47 for cases and 394 for non-cases household contacts.
‡Laboratory evidence of SARS-CoV-2 infection included positive molecular or antigen testing and was self-reported.
§Excludes any additional tests obtained after the first positive SARS-CoV-2 molecular or antigen test.

The SAR was 9% (48/526; 95% CI, 7%–12%). Among household contacts who reported molecular or antigen SARS-CoV-2 testing, the SAR was 12% (46/377; 95% CI, 9%–16%). Five (10%) of 48 secondary cases compared with 130 (33%) of 398 non-case household contacts reported potential community exposures. Secondary cases occurred in 35 (18%) of 194 households (Figure 2); among households with secondary cases, the SAR was 45% (48/107; 95% CI, 36–54%).

**Figure 2.**
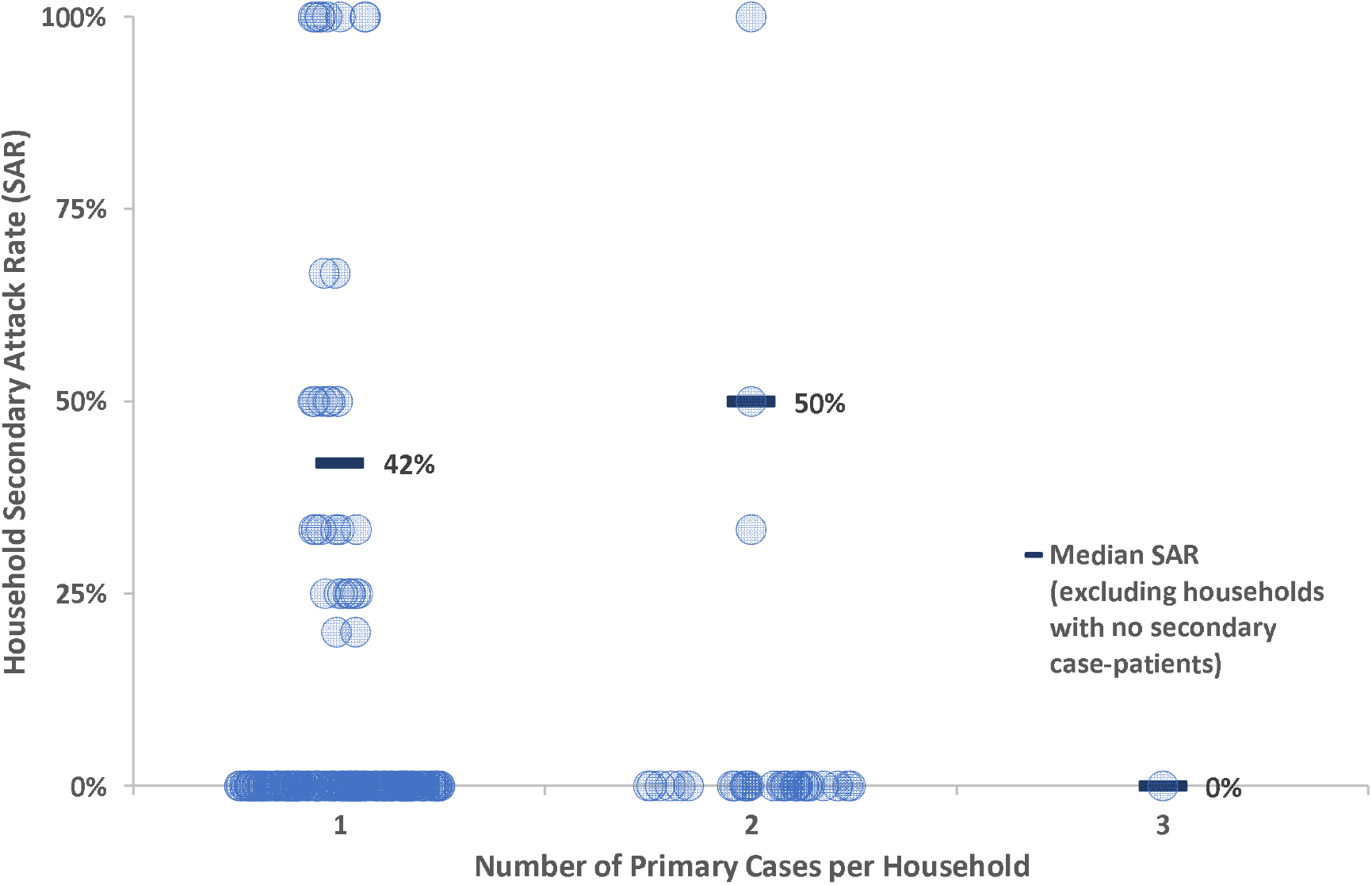
Scatter plot of household secondary attack rates (SAR) by the number of primary cases per household and the median SAR, excluding households with no secondary cases, by the number of primary cases per household

## Discussion

Following widespread transmission among camp attendees at an overnight camp,^12^ we found that children and adolescents transmitted SARS-CoV-2 to pediatric and adult household contacts, consistent with transmission dynamics of other viral respiratory diseases.^14^ Transmission from children to adults resulted in 10% of the adult secondary cases requiring hospitalization.

Previous household transmission investigations found an overall SAR ranging from 11%–32%, although these studies focused on adult primary cases and tested all household contacts for SARS-CoV-2.^15-20^ Our finding of a 12% SAR among tested household contacts is in the lower end of the SAR range reported by other studies. However, due to a known camp exposure, many primary cases isolated or wore masks upon returning home. Additionally, not all household contacts were systematically tested. Furthermore, 20% of the primary cases had symptom onset while at camp. This is notable given laboratory and epidemiologic evidence indicating that transmission risk is highest during the pre-symptomatic and early onset period of illness.^21,22^ In comparison, an investigation of childcare facility-associated outbreaks with pediatric primary cases noted a 26% SAR among contacts.^23^

This investigation includes a large cohort of children and adolescents identified as primary cases in their households and adds valuable evidence for SARS-CoV-2 transmission from children and adolescents. However, our findings are subject to at least three limitations. First, this was a retrospective observational study with selection and recall bias. Second, the participating household contacts were predominantly non-Hispanic White with a college degree and therefore not representative of the general population. Third, differential misclassification leading to SAR underestimation might have occurred as not all household contacts were tested for SARS-CoV-2, test results were self-reported, and many contacts were tested only once and could have been tested too early. Alternatively, SAR could have been overestimated due to inability to distinguish household or community SARS-CoV-2 infection source, and between secondary and tertiary household transmission.

In this investigation, school-aged children and adolescents with COVID-19 transmitted SARS-CoV-2 to other children and adults in the household setting, with 10% of secondary adult cases requiring hospitalization. These findings highlight the importance of implementing effective public health guidelines to prevent SARS-CoV-2 transmission in all settings, including settings with children. Children and adolescents should remain at home, ≥6 feet apart from contacts, and have a separate sleeping space and bathroom following a known COVID-19 exposure or diagnosis. In communities with active SARS-CoV-2 spread, and particularly in congregate settings, children and adolescents should wear masks if safe to do so and maintain at least six feet distance from others to prevent SARS-CoV-2 transmission.^24,25^

## Data Availability

Deidentified data to reproduce results will be available upon peer-reviewed publication and request.

## Acknowledgements

*Camp attendees and their household contacts*

*Georgia Department of Public Health:* Luke Baertlein, Tiffany Baird, Aaron Blakney, Tom Campbell, Alicia Dunajcik, Amit Eichenbaum, Amanda Feldpausch, Pamela Logan, Amanda Mohammed, Stephanie O’Conner, Haley Putnam, Zoe Schneider, Brandon Shih, Kat Topf, and Bill Williamson

*Centers for Disease Control and Prevention:* Ramika Archibald, Elizabeth Dietrich, Kathy Fowler, Leah Graziano, Chad Heilig, Margaret Honein, Mark Johnson, Kelsey McDavid, Robert Montierth, Anupama Shankar, Robert Slaughter

*State health departments:* Alabama Department of Public Health, Arkansas Department of Health, Colorado Department of Public Health and Environment, Florida Department of Health, Maryland Department of Health, North Carolina Division of Public Health, South Carolina Department of Health and Environmental Control, Tennessee Department of Health, Texas Department of State Health Services

